# Predicting the epidemiological effects in the United Kingdom of moving from PCV13 to PCV15 in the routine pediatric 1+1 vaccination schedule

**DOI:** 10.1101/2025.03.19.25324262

**Authors:** Rachel J Oidtman, Natalie Banniettis, Jessica Weaver, Ian R Matthews, Dionysios Ntais, Giulio Meleleo, Tufail M Malik, John C Lang, Oluwaseun Sharomi

## Abstract

**Introduction:** In 2020 the UK National Immunization Programme reduced the pediatric dosing schedule for pneumococcal vaccination with PCV13 from 2 doses in infancy followed by a toddler dose (2+1 schedule) to 1 dose in infancy followed by a toddler dose (1+1 schedule). Real world data on vaccine effectiveness (VE) against invasive pneumococcal disease (IPD) under the reduced dosing schedule is not available, nor is data on VE associated with higher valency vaccines, including PCV15. This analysis investigates changes in projected disease outcomes associated with potential reductions in VE against IPD with PCV13 and PCV15 under the 1+1 reduced dosing schedule.

**Methods:** A previously published dynamic transmission model was employed to project population-level IPD incidence under 1+1 pneumococcal vaccination dosing of children <2 years old with two different vaccine formulations, with maintenance of current adult and at-risk vaccination programs, under a range of assumptions regarding reduction of VE against IPD, over a 20-year timeline following the switch to the 1+1 schedule. A probabilistic sensitivity analysis was performed to evaluate the sensitivity of model outcomes to potential changes in VE against IPD.

**Results:** Relative to 2019 estimates, overall IPD incidence was predicted to increase over the time horizon for both PCV13 and PCV15 programs, however vaccination with PCV15 was estimated to lead to a smaller increase over all age groups than vaccination with PCV13. Both vaccine formulations led to projected decreases in IPD attributed to common PCV13/PCV15 serotypes in children <2 years old. Sensitivity analyses reflect results were robust to potential changes in VE against IPD.

**Conclusions:** The current analysis predicts switching the routinely administered pediatric pneumococcal vaccine from PCV13 to PCV15 within the UK’s 1+1 dosing schedule would not only reduce IPD in the pediatric population but would also lead to population-level reductions in IPD due to indirect protection.

## INTRODUCTION

Pneumococcal disease (PD), caused by *Streptococcus pneumoniae*, leads to significant morbidity and mortality, especially in young children.^1^ Preventative pneumococcal conjugate vaccines (PCVs) targeting clinically relevant serotypes are recommended for routine administration in early childhood. PCV7 was initially licensed 25 years ago as a 4-dose schedule, with 3 doses in infancy followed by a toddler dose (3+1).^1^ Based on immunogenicity data supporting reduced dosing schedules, coupled with economic pressures and crowded childhood immunization schedules, the United Kingdom (UK) became the first country to adopt PCV7 in a 3-dose (2+1) schedule in their national immunization programme (NIP) in 2006.^2,3^ In the adult population, PPSV23 has been widely used for adults aged ≥65-years-old since 2003.^4^

Before the introduction of PCVs in the UK, overall invasive pneumococcal disease (IPD) incidence was 14.8 per 100,000, with the PCV7 serotypes accounting for approximately half of the IPD disease burden.^4^ By 2010, the overall incidence of IPD had dropped to 10.1 cases per 100,000, with PCV7 serotypes comprising 14% of IPD.^4^ However, a relative increase in the proportion of non-vaccine type (NVT) PD led to the adoption of PCV13 in 2010, which covered 58% of the disease burden at that time.^4^ By 2019-2020, overall disease incidence reached a low of 9.4 per 100,000 and NVT IPD comprised about 81% of the disease burden.^1^ The reduction in overall IPD, with NVT driving the residual burden of disease, motivated two changes for the pediatric PCV NIP – the move to a 1+1 dosing schedule, and the interest in switching to a higher valency PCV. These changes were further supported by several factors: (1) high vaccine coverage rates leading to good control of vaccine type (VT) PD;^3^ (2) a 2018 randomized control study by Goldblatt et al which demonstrated similar immune responses to PCV13 in 2+1 and 1+1 dosing schedules;^2^ and (3) a 2019 modeling study by Choi et al projecting the incidence of IPD and non-bacteremic pneumococcal pneumonia (NBPP) would not be affected by a reduction in dosing schedule.^5^

The decision to move to a 1+1 dosing schedule was not without risk, as fewer doses in infancy, when the immune system is immature, may lead to subpar protection and breakthrough disease. A separate modeling study conducted by Wasserman et al explored the projected potential effects of switching to a 1+1 PCV13 dosing schedule in the UK.^6^ This study estimated the reduction of the number of infant doses was projected to lead to an increase in IPD cases over a 10-year time horizon, with the greatest increases occurring in infants and in older adults, also resulting in an increase in deaths.^6^ This finding was consistent with the results of the randomized control study by Goldblatt et al which supported the switch to a 1+1 dosing schedule while also noting this potential risk.^2^

In January of 2020, the 1+1 reduced dosing schedule was implemented in the UK, with the infant dose administered at 12 weeks of age and the toddler dose at 1 year.^7^ Contagion control measures for the COVID-19 pandemic were implemented soon after this switch, rendering it difficult to estimate the effects of this policy change due to the overall reduction of PD. A real-world observational study, published in 2024, indicated that the overall IPD incidence in 2022/23 had decreased in the years since implementation of the 1+1 dosing schedule (which included the period when COVID-19 pandemic measures were in effect).^1^ However, VT IPD incidence increased over this period, which may hint at the fact that the immune response for a 1+1 schedule is insufficient to maintain prior levels of control of VT IPD, though it is difficult to draw conclusions given the confounding effects of COVID during this timeframe.^1^

The percentages of PD due to VT and NVT in the UK are still substantial and warrant the consideration of higher valency PCVs in the pediatric NIP. Due to the reduction of serotype-specific immunogenicity as more serotypes are added to a PCV (“immunogenicity-creep”), there is risk of reduced effectiveness of higher valency PCVs.^8,9^ Nevertheless, the replacement of PCV7 with the higher-valency PCV13 in the UK did not lead to an increase in VT disease.^4^ Two new expanded valency PCVs were recently licensed in the UK – PCV15 (Vaxneuvance^TM^, Merck & Co., Inc., Rahway, NJ, USA) and PCV20 (Prevnar 20^TM^, Wyeth Pharmaceuticals LLC, a subsidiary of Pfizer, Inc.) – which provide protection against additional NVT serotypes.^1^ The clinical impact of these new PCVs and their potential for immunogenicity-creep expected in 1+1 dosing remains to be seen.

The confluence of information from immunogenicity studies on reduced dosing schedules, epidemiological data, transmission models, and the impact of the COVID 19 pandemic on the epidemiology of infectious diseases paints a confusing picture of the potential effectiveness of a 1+1 dosing schedule for pediatric pneumococcal vaccination in the UK with PCV13, and potentially with higher valent PCVs. This analysis employs a previously described and calibrated model to explore the potential impact of reduced vaccine effectiveness (VE) against disease resulting from a reduced dosing schedule, with pediatric vaccines PCV13 and PCV15.

## METHODS

### Model overview

The dynamic transmission model used in this analysis was a published deterministic, age-structured, population-level model, accounting for (1) demographic components, including births, deaths, and aging, and (2) carriage transmission dynamics in the presence of historical vaccine introduction.^10,11^ The demographic model assumed a constant population size and included age structure in the form of four age groups: <2-, 2-4-, 5-64-, and ≥65-year-olds.^12,13^ Mixing among populations of different ages is particularly relevant for *S. pneumoniae* transmission and was accounted for with a previously published mixing matrix.^14^

The epidemiological model was based on a system of ordinary differential equations and distinguished between pneumococcal carriage (single, double, and triple) and disease, as well as vaccination status. The progression from pneumococcal carriage to pneumococcal disease was modeled through an age-, serotype class-(STC), and vaccine-status-specific relationship, wherein disease occurs in some fraction of carriage episodes (Supplementary Figure S1). Given the relative infrequency of transmission from adults to children, the simplifying assumption was made that no transmission occurred from adults to children.^15^ For the purpose of this model, pediatric age strata were comprised of <2- and 2-4-year-olds and adult age strata were comprised of 5-64- and ≥65-year-olds.

A complete description of the model, inputs, parameter estimates, and calibration results was provided in Oidtman et al. 2025.^11^ In brief, the model was calibrated using the Nelder-Mead simplex method implemented in the NMinimize function in Mathematica 13.3.1 (Wolfram Research, Champaign, IL) to minimize a weighted sum of squared errors objective function over the annual age- and STC-specific IPD incidence data.^16^ During the calibration, four sets of age- and STC-specific parameters were estimated: vaccine efficacy against carriage, carriage acquisition rate given contact, invasiveness (case-to-carrier ratio), and pairwise competition between STCs.^11^ For the competition parameters, historical replacement dynamics were used to calibrate the specific STC-STC combinations. In the absence of observed data on replacement dynamics, such as is the case between historical novel VTs in PCV13 and the novel VTs in PCV15, there was assumed to be no explicit competition.

### Inputs and data sources

Annual age- and serotype-specific IPD incidence data for 2000-2019 were provided by the UK Health Security Agency (HSA)^4^ and were used as a model calibration target for the pre-PCV NIP era steady state (2000-2005) and the vaccine period (2006-2019). As the model is driven by carriage dynamics, *S. pneumoniae* carriage data from 2006 were used as a baseline target for the pre-PCV NIP era.^17,18^ Serotype-specific IPD and carriage data were aggregated into 11 STCs based on the inclusion of different serotypes in different vaccines to calibrate to historical data (Table 1).^19,20^

**Table 1.** Serotype classes (STCs) with respective serotypes (STs) included in each class. Shading indicates inclusion in different vaccines. The vaccination group indicates how serotype classes were grouped for visualization purposes. Note that serotype 3 was included in a stand-alone STC due to evidence of limited immunogenicity response from vaccines.^21^

| Serotype class (STC) | Serotypes included in class | Vaccination Group | PCV7 | PCV13 | PCV15 |
| --- | --- | --- | --- | --- | --- |
| 1 | 4, 6B, 9V, 14, 18C, 19F, 23F | Common PCV13/PCV15 STs | - | - | - |
| 2 | 1, 5 | Common PCV13/PCV15 STs |  | - | - |
| 3 | 3 | Common PCV13/PCV15 STs |  | - | - |
| 4 | 7F, 19A | Common PCV13/PCV15 STs |  | - | - |
| 5 | 6A, 6C<sup>A</sup> | Common PCV13/PCV15 STs |  | - | - |
| 6 | 22F, 33F | PCV15 Unique STs |  |  | - |
| 7 | 9N, 17F, 20 | NVTs |  |  |  |
| 8 | 8, 10A, 11A, 12F | NVTs |  |  |  |
| 9 | 15B | NVTs |  |  |  |
| 10 | 15A, 15C, 16F, 23A, 23B, 24F, 31, 35B | NVTs |  |  |  |
| 11 | NVTs<sup>B</sup> | NVTs |  |  |  |

NVT = non-vaccine type; ST = serotype.
<sup>A</sup> ST 6C is included in STC 6 due to assumed cross protection with serotype 6A.
<sup>B</sup> To simplify the model, the number of STCs was reduced by classifying ST 2 as an NVT. This assumption is anticipated to have a minimal impact on model predictions due to the low prevalence of ST 2.

Serotype and age-based data on VE against IPD were available for PCV13 ^22^ and PPSV23.^23^ Given the lack of real-world evidence for VE against IPD for novel PCV15 serotypes, VE was estimated as the weighted average (weighted by the number of STs in each PCV grouping) from prior vaccine-specific estimates (Supplementary Tables S1-S2). Base case values for VE against IPD (Table 3) were calculated by applying a 12.8% average reduction in PCV13 VE under a 2+1 schedule, based on an observational study by Savulescu et al;^22^ additional details are provided in the Sensitivity analysis subsection. Data on serotype-specific carriage clearance rates (i.e., the inverse of the duration of carriage) were available from studies from multiple countries and settings,^24–36^ which were averaged over serotype and age to obtain aggregate inputs into the model (more details available in Tables S3-S5 in Oidtman et al. 2025 ^11^). Data on vaccination coverage rates by age were available from the HSA (Supplementary Tables S3-S5).^37^

### Projection scenarios

The calibrated model was used to evaluate the impact of implementing a 1+1 dosing schedule among children <2 years old with either PCV13 or PCV15 beginning in 2020. Both scenarios assumed a continuation of PPSV23 in older adult and risk group populations. In both pediatric and adult populations, it was assumed VCR levels from 2019 remained constant into the projected 20-year time horizon (Supplementary Tables S3-S5). Reductions in pneumococcal disease or contact patterns arising from protection measures enacted during the COVID-19 pandemic were not accounted for in these projections. Model projection scenarios are summarized in Table 2.

**Table 2.**
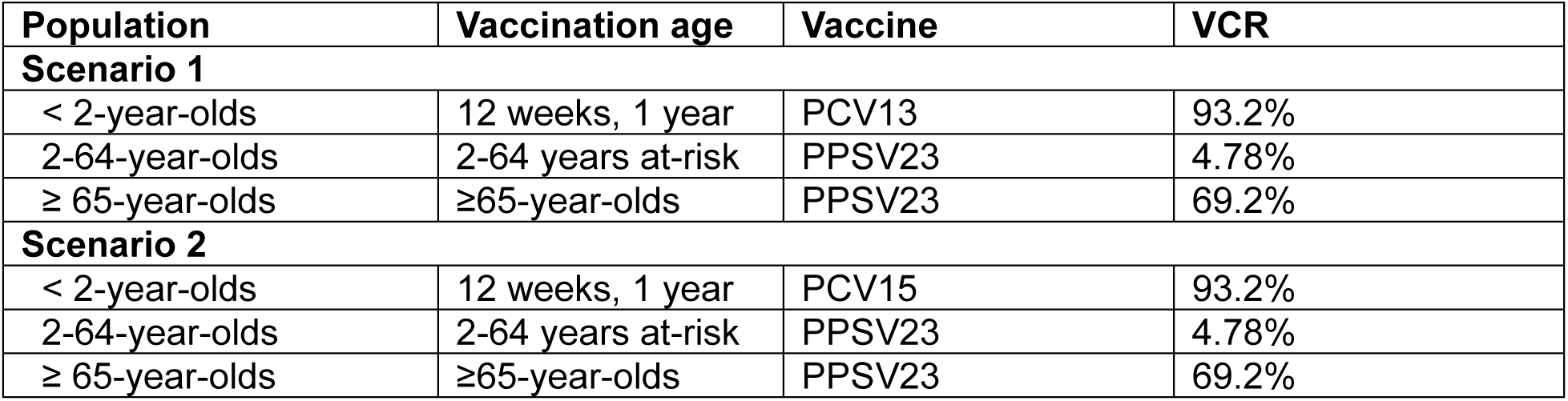
Projected vaccination scenarios for PCV13 and PCV15 in a 1+1 dosing schedule.

| Population | Vaccination age | Vaccine | VCR |
| --- | --- | --- | --- |
| **Scenario 1** |  |  |  |
| < 2-year-olds | 12 weeks, 1 year | PCV13 | 93.2% |
| 2-64-year-olds | 2-64 years at-risk | PPSV23 | 4.78% |
| ≥ 65-year-olds | ≥65-year-olds | PPSV23 | 69.2% |
| **Scenario 2** |  |  |  |
| < 2-year-olds | 12 weeks, 1 year | PCV15 | 93.2% |
| 2-64-year-olds | 2-64 years at-risk | PPSV23 | 4.78% |
| ≥ 65-year-olds | ≥65-year-olds | PPSV23 | 69.2% |

### Sensitivity analysis

To explore uncertainty in the potential reductions in VE against IPD associated with a reduced pediatric dosing schedule, a probabilistic sensitivity analysis (PSA) was performed which varied the vaccine effectiveness against IPD in both PCV13 and PCV15. This analysis did not consider potential reductions in vaccine efficacy against carriage.

Potential reductions in VE against IPD resulting from the adoption of a reduced dosing schedule were informed by the estimated 12.8% average reduction in PCV13 VE when shifting from a 3+1 schedule to a 2+1 schedule, based on an observational study by Savulescu et al of PCV13 VE against IPD in pediatric populations in Europe.^22^ The range in VE values for the PSA was defined as follows: the upper limit was set to equivalency, such that the VE would be equivalent in a 1+1 and 2+1 dosing schedule; the mean value was set to the average 12.8% reduction following Savulescu et al.;^22^ and the lower limit was set to a maximum reduction informed by doubling the average reduction (Table 3).

**Table 3.** Vaccine efficacy values used for the probabilistic sensitivity analysis. 12.8% average reduction corresponds to the reduction from 0.897 (3+1) to 0.782 (2+1) as described in Savulescu et al.^22^

| STC (STs) | Serotype grouping | Maximum VE<sup>A</sup> | Base case VE<sup>B</sup> | Minimum VE<sup>C</sup> |
| --- | --- | --- | --- | --- |
| 1 (4, 6B, 9V, 14, 18C, 19F, 23F) | Common PCV13/PCV15 STs | 0.96 | 0.84 | 0.71 |
| 2 (1, 5) | Common PCV13/PCV15 STs | 0.84 | 0.74 | 0.63 |
| 3 (3) | Common PCV13/PCV15 STs | 0.41 | 0.36 | 0.31 |
| 4 (7F, 19A) | Common PCV13/PCV15 STs | 0.93 | 0.82 | 0.69 |
| 5 (6A, 6C) | Common PCV13/PCV15 STs | 0.96 | 0.84 | 0.71 |
| 6 (22F, 33F) | PCV15 Unique STs | 0.94 | 0.83 | 0.70 |

STC – serotype class; ST – serotype; VE – vaccine effectiveness.
<sup>A</sup> Maximum VE was assumed to be equal to VE for the 2+1 vaccine schedule.
<sup>B</sup> Base case VE was assumed to be 12.8% lower than VE for the 2+1 vaccine schedule.
<sup>C</sup> Minimum VE was assumed to be equal to $2 \times 12.8\% = 25.6\%$ lower than VE for the 2+1 vaccine schedule.
<sup>D</sup> VE for STC7-11 were assumed to be 0 for PCV7, PCV13, and PCV15 vaccines.

The VEs were drawn from beta distributions with ranges approximately equal to those described in Table 4 and parameters were varied in a Latin Hypercube Sampling method with a total of 100 random samples.^38^ The PSA did not consider differences in VE by vaccine, for example a reduced VE in PCV7 STs was applied equally to both PCV13 and PCV15.

**Table 4.** Projected IPD incidence (per 100,000) by serotype grouping in <2-year-olds. IPD incidence at the start of the projection (0 years) and at the end of the 20-year time horizon, with administration of either PCV13 or PCV15 in the <2-year-old population. The mean projections of IPD (over 100 Latin hypercube samples), percent change from start of projections, and the 90% intervals (5% and 95% quantiles) are included. NA values indicate the same value across all PSA realizations and therefore no informative interval. NA – not applicable

| Serotype grouping | Initial IPD incidence at 0 years | Average IPD incidence at 20 years (change versus initial incidence) | 90% interval for IPD incidence at 20 years |
| --- | --- | --- | --- |
| **Pediatric vaccination with PCV13** |  |  |  |
| Common PCV13 / PCV15 serotypes | 4.04 | 3.23 (-20.0%) | (2.78, 3.74) |
| PCV15 unique serotypes | 1.75 | 1.86 (+6.3%) | NA |
| Serotypes not included in PCV13 | 10.63 | 13.08 (+23.0%) | NA |
| Serotypes not included in PCV15 | 8.88 | 11.22 (+26.4%) | NA |
| Overall | 14.67 | 16.30 (+11.1%) | (15.90, 16.80) |
| **Pediatric vaccination with PCV15** |  |  |  |
| Common PCV13 / PCV15 serotypes | 4.04 | 3.45 (-14.6%) | (2.96, 4.01) |
| PCV15 unique serotypes | 1.75 | 0.12 (-93.1%) | (0.10, 0.15) |
| Serotypes not included in PCV13 | 10.63 | 11.73 (+10.3%) | (11.71, 11.76) |
| Serotypes not included in PCV15 | 8.88 | 11.61 (+30.7%) | NA |
| Overall | 14.67 | 15.18 (+3.5%) | (14.70, 15.80) |

## RESULTS

### Model calibration

The model was able to closely reproduce historically observed IPD dynamics, including the decline in incidence of VT IPD following the introduction of PCV7 and PCV13 and the subsequent increase of NVT IPD due to serotype replacement. In the <2- and 2-4-year-old populations, there were some observed dynamics that the model was not able to completely reproduce, such as the large increases of non-PCV15 serotypes beginning in 2014-2015 (Figure S2).

The root mean squared error (RMSE) was used post hoc to compare model predictions to surveillance data by age, year, and STC, wherein an RMSE closer to zero indicates a better fit while a higher value indicates more error. The RMSE values were 1.47, 0.597, 0.312, and 1.29 for the <2-, 2-4-, 5-64-, and ≥65-year-old age groups (Figure S3). Across all age groups, the RMSE was 1.03. A complete description of the model fitting results and parameter estimates are available in Oidtman et al. 2025.^11^

### Projections of IPD in children less than two years old

For both PCV13 and PCV15, the model projected an increase in overall IPD incidence in children less than two years old over a 20-year time horizon as a consequence for switching from a 2+1 to a 1+1 dosing schedule in 2020 (Figure 1). Model outcomes for PCV13 and PCV15 reflect an overlap in projected IPD incidence for shared STCs, with a 14.6%-20.0% decline in the incidence of serotypes covered by both PCV13 and PCV15 over the 20-year time horizon following the switch to the reduced dosing schedule (Figure 1a and 1b, Table 4). IPD incidence due to PCV15-unique STCs declined by 93.1% over the time horizon with implementation of PCV15 and increased by 6.3% with continued PCV13 vaccination (Figure 1c and 1d, Table 4). IPD incidence due to serotypes not included in PCV13 increased by 10.3% and 23.0% (Figure 1e and 1f), and IPD incidence due to serotypes not included in PCV15 increased by 30.7% and 26.4% for PCV15 and PCV13 scenarios, respectively (Figure 1g and 1h, Table 4). The absolute increase in overall IPD over the time horizon was smaller for PCV15 than for PCV13 (3.5% vs 11.1%) (Figure 1i and 1j, Table 4). Sensitivity analyses reflect results were robust to potential changes in VE against IPD.

**Figure 1.**
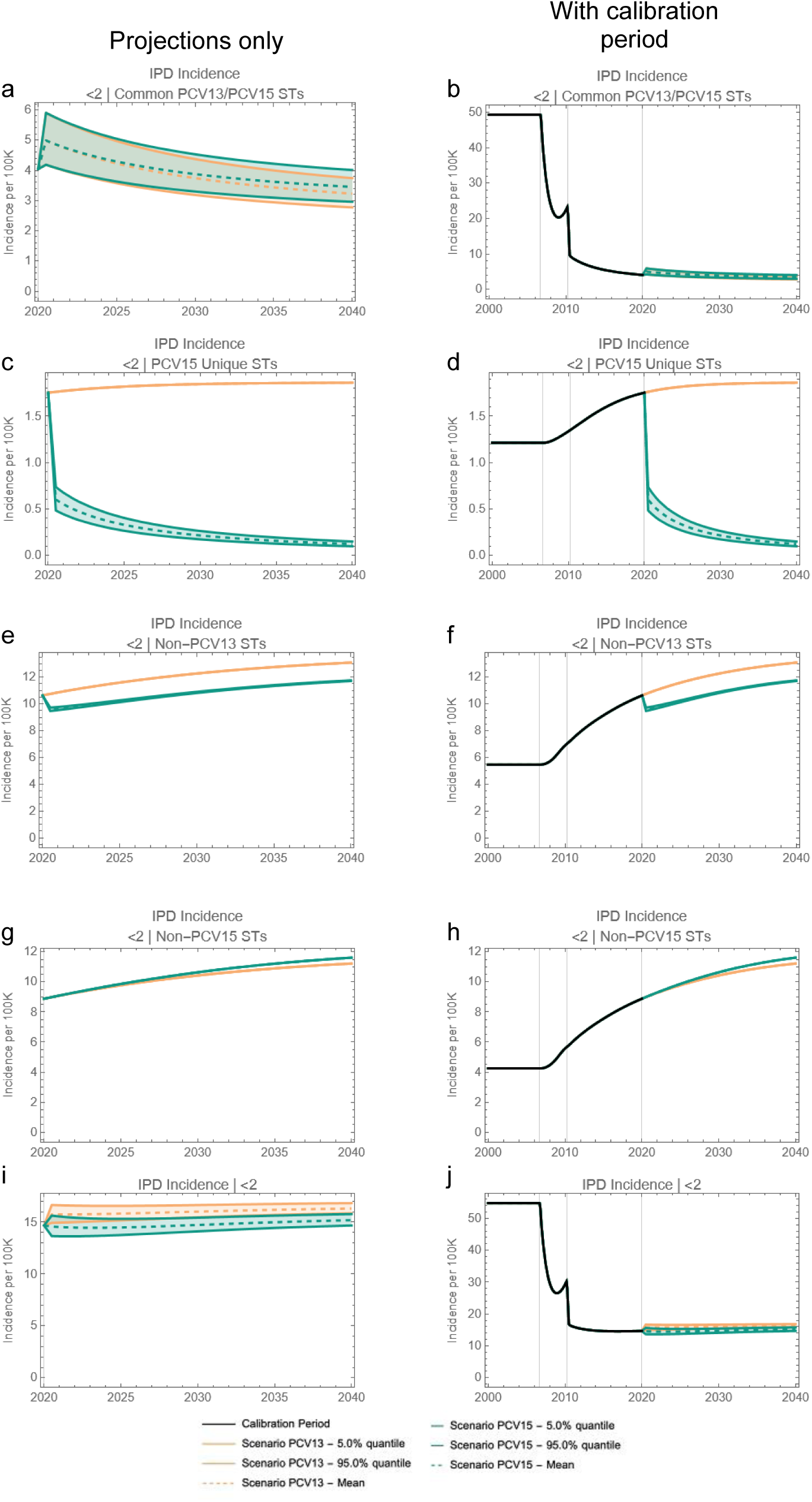
20-year projections of IPD incidence in <2-year-olds by serotype grouping. The two columns showcase the same results; the left column focuses on the projection period only while the right column includes the calibration period. The first three rows illustrate IPD by ST grouping and the fourth row illustrates overall IPD incidence in <2-year-olds. ST – Serotype.

### Population-level projections of IPD

At the population level, projected overall IPD incidence outcomes for PCV13 and PCV15 overlapped for children aged 2-4-years-old over the duration of the 20-year time horizon (Figure 2a and 2b). Further, the model projected a 4.2% decrease in IPD incidence in the 2-4-year-old age group following the introduction of PCV15, compared with negligible change in IPD incidence for this same age group with implementation of PCV13 (Table 5). The model projected a smaller increase in IPD incidence with the introduction of PCV15 when compared with PCV13 for the <2-(3.5% increase vs 11.2% increase, Table 4) and 5-64-year-old (5.1% increase vs 7.8% increase) age groups, as well as for the overall population (0.6% increase vs 7.1% increase, Figure 3 and Table 5). As seen with children <2 years of age, sensitivity analyses reflected projected IPD incidence results for all modeled age groups were robust to potential changes in VE against IPD.

**Figure 2.**
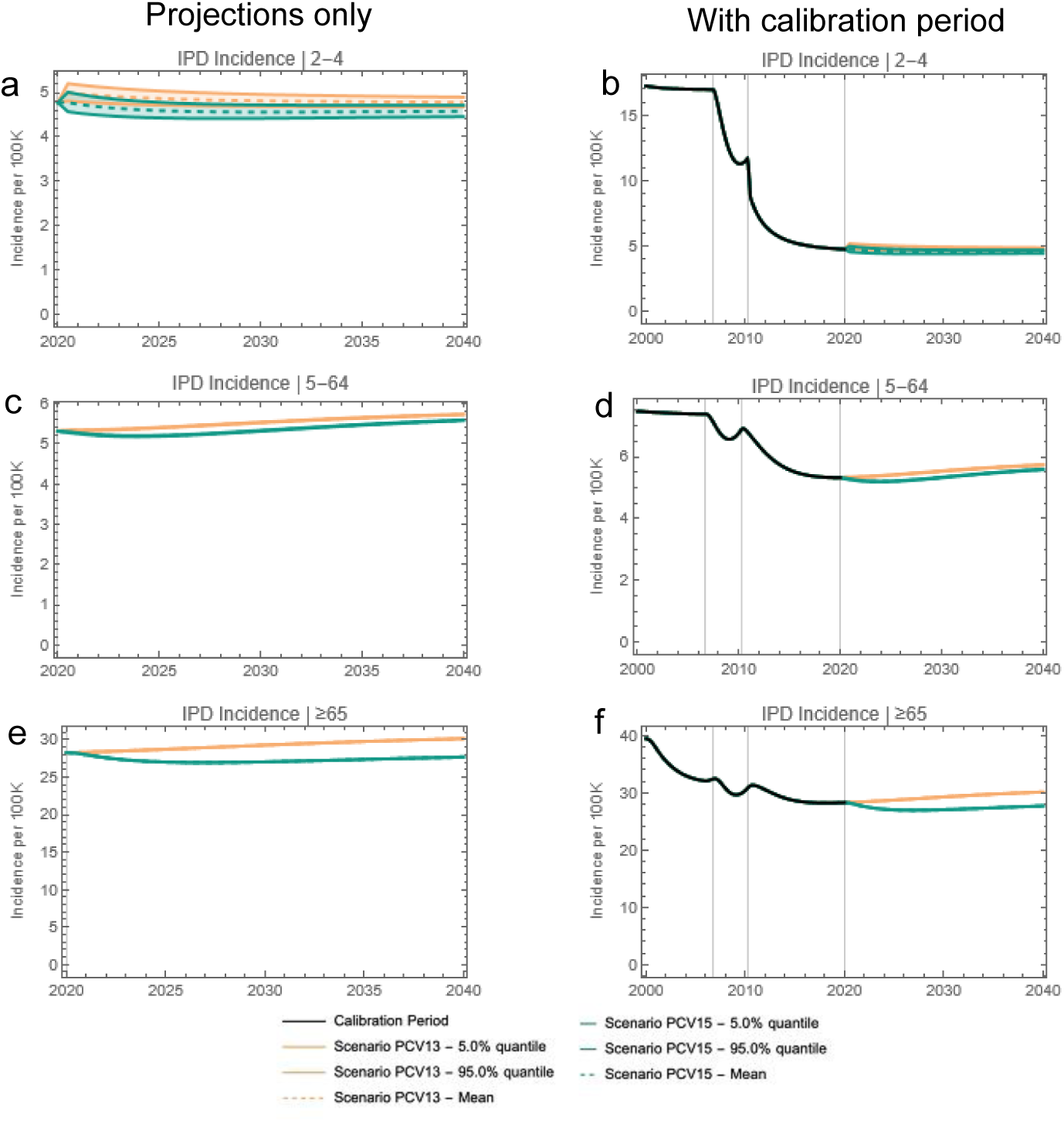
20-year projections of IPD in 2-4-, 5-64-, and ≥65-year-olds. The two columns showcase the same results; the left column focuses on the projection period only while the right column includes the calibration period. Each row represents a different age group.

**Figure 3.**
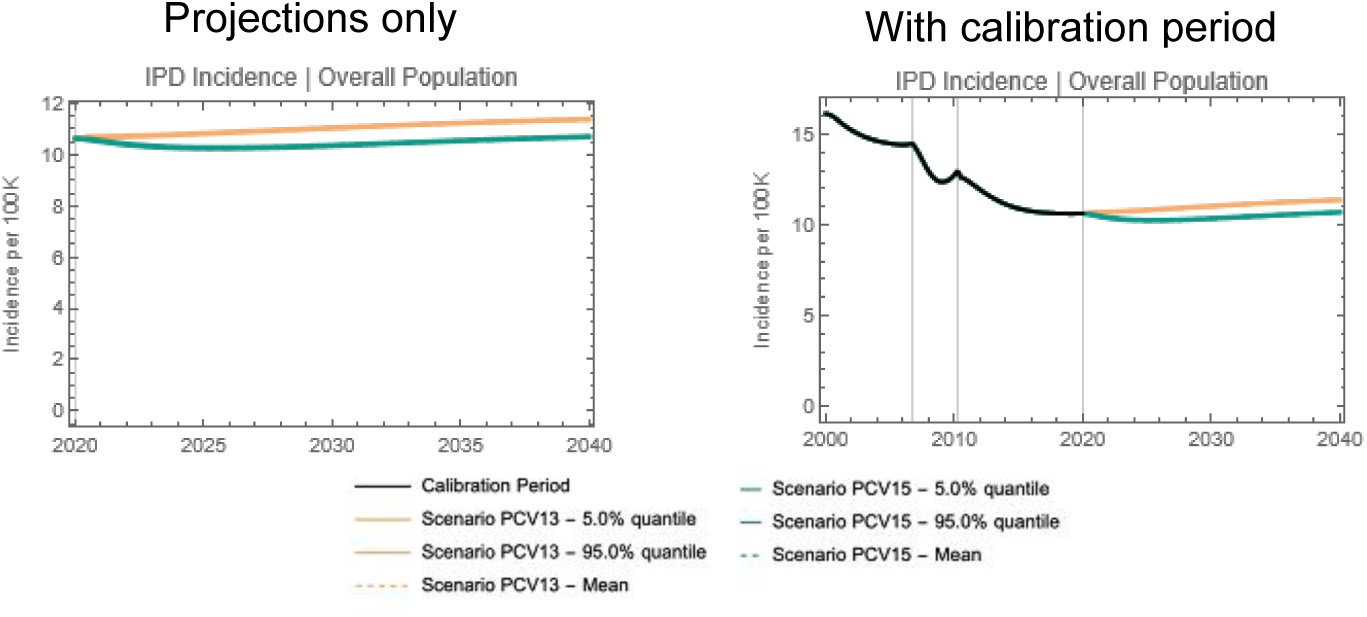
20-year projections of IPD in the entire population. The two columns showcase the same results; the left column focuses on the projection period only while the right column includes the calibration period.

**Table 5.**
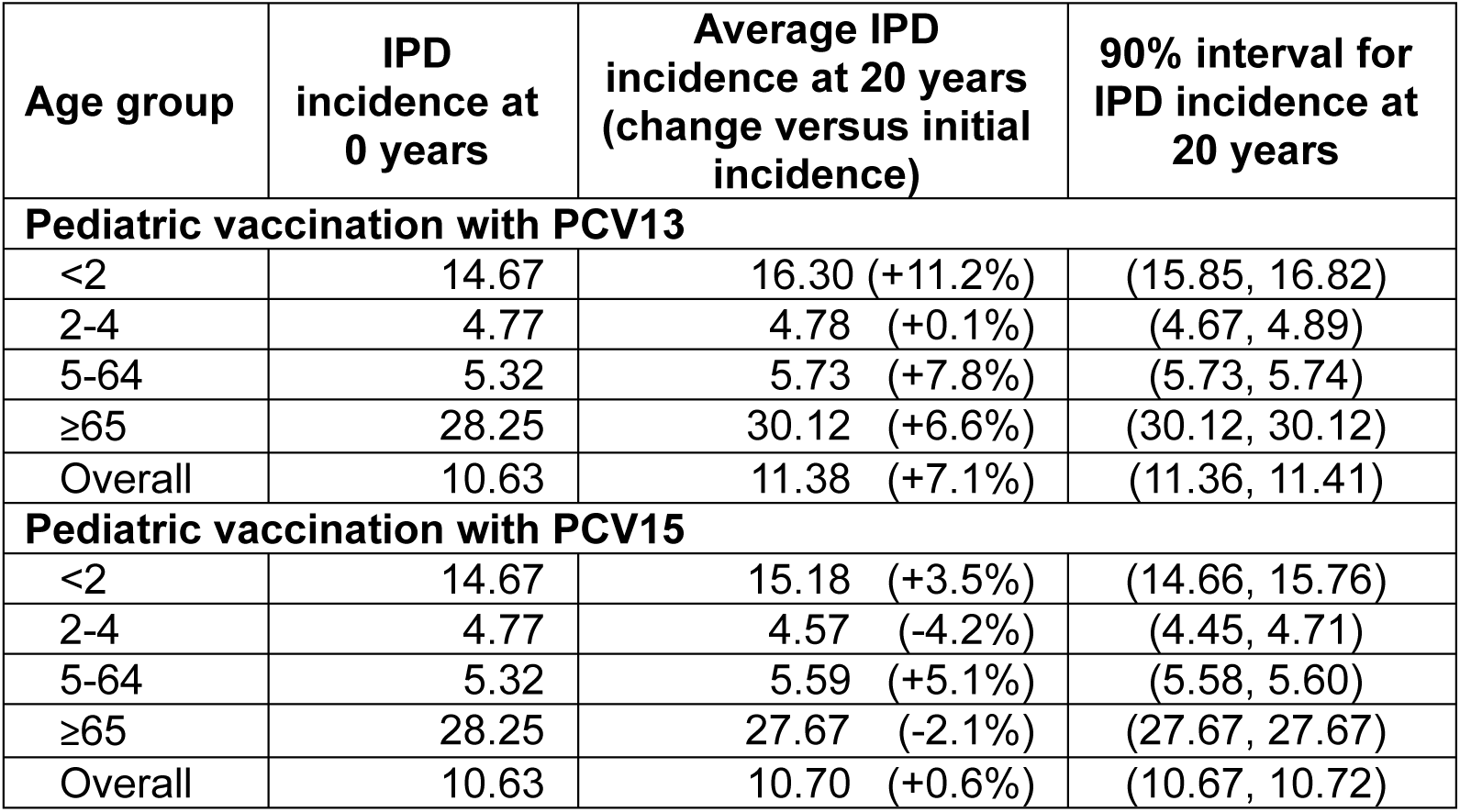
Projected IPD incidence (per 100,000) by age. IPD incidence at the start of the projection (0 years) and at the end of the 20-year time horizon, with administration of either PCV13 or PCV15 in the <2-year-old population. The mean projections of IPD, percent change from start of projections, and the 90% confidence intervals (5% and 95% quantiles) are included.

## DISCUSSION

Despite the inclusion of effective PCVs in pediatric vaccination programs, residual pneumococcal disease remains a public health matter in the UK and throughout the world. Here, a previously described and calibrated dynamic transmission model was used to investigate the clinical outcomes resulting from potential reductions in VE with two PCVs (PCV13 and PCV15) when routinely administered in a reduced pediatric dosing schedule (1+1) in the UK.^10,11^ Model projections predict the use of PCV15 would significantly reduce IPD incidence both in children <2 years of age, due to direct protection, and in older children and adults, due to indirect protection, with respect to continued use of PCV13. These effects were consistent regardless of potential changes in VE against disease.

Reductions in IPD in children <2 years old attributed to the serotypes unique to PCV15 (22F and 33F) drove PCV15 to avert more disease than PCV13 in model projections. While both PCV13 and PCV15 led to projected decreases in IPD attributed to serotypes covered by both PCV13 and PCV15 in this age group, PCV15 led to a smaller projected decrease when compared with PCV13. Marginal increases in non-PCV15 serotypes were projected.

Nevertheless, these differences were insufficient to counteract the overall estimated clinical benefits of pediatric PCV15 vaccination observed in the <2-year-old age group and in the population as a whole. With either vaccine, relative to current 2019 estimates of IPD incidence, overall IPD incidence was predicted to increase over the 20-year time horizon, though the projected overall increase was predicted to be smaller with implementation of PCV15 than with PCV13 vaccination.

In contrast, a recent model by Choi et al evaluated the implications of introducing higher valency pediatric vaccines in the UK via the 1+1 schedule and concluded the introduction of PCV15 could result in increased IPD relative to the standard of care of PCV13.^7^ These findings were in part a function of this study’s projection assumption on serotype replacement, wherein a decline in the carriage of a VT ST would be met with 100% replacement of carriage with a NVT ST, although this assumption is not substantiated by historical data to date. While it is necessary to make assumptions regarding the level of replacement expected upon the introduction of higher valency vaccines, assuming 100% replacement may lead to unreasonably high predictions of disease incidence of NVT STs. In contrast, the current analysis estimated competition between PCV13 and non-PCV13 STs through model calibration. However, there were insufficient data to estimate competition parameters between PCV15 and non-PCV15 STs, thus minimal competition between PCV15 and non-PCV15 STs was assumed. In addition, the Choi et al analysis implemented two separate calibration models for model fitting to historical data, which has the potential to introduce bias and confounding effects due to incompatibilities between the two models.^7^ The current analysis employed a single model for calibration and fitting, to ensure compatibility and consistency.

Several simplifying assumptions were made in the current analysis due to model complexity and limited availability of data. Potential reduction in VE against disease resulting from a change to a reduced pediatric vaccination schedule was considered, but there may also be reduced duration of protection and reduced VE against carriage.^6^ Reductions in VE against carriage may lead to an increase in VT carriage, which could then increase VT-IPD in pediatric populations due to direct effects and adult populations due to indirect effects. In addition, consistent with previous modeling studies,^7^ we did not consider the possibility of immunogenicity creep with the introduction of PCV15. This will be an important area of future research as real-world data become available for expanded valency PCVs.

There were several limitations to this analysis. First, the model is very sensitive to estimates of calibrated parameters, including VE against carriage. Although this model considered a range of potential reductions in VE against disease that could occur resulting from a change to a reduced dosing schedule, it could not consider potential changes in VE against carriage without conducting a more in-depth parameter identifiability analysis. Second, the PSA only considered the VE against disease after the toddler dose (i.e., at the completion of the 1+1 dosing schedule). VE in the first year of life (i.e., the VE after receiving the single infant dose in the 1+1 schedule) may be more critical to evaluate given the vulnerability of infants to IPD.^5^ However, since the model is not dose dependent, it could not explicitly predict the effects of two infant doses (in a 2+1 schedule) compared to a single infant dose (in a 1+1 schedule). Modeling dose dependent VEs and the potential for breakthrough IPD in infants with a reduced dosing schedule will be an important avenue for future research. Third, as there are limited data on real-world estimates of VEs against disease in a 1+1 dosing schedule, in part due to the COVID-19 pandemic coinciding with the start of the reduced dosing schedule in the UK, this PSA was instead informed by average changes when moving from a 3+1 to 2+1 dosing schedule.^22^ As more estimates of VE against disease and carriage become available, it will be important to incorporate these into model-based predictions. Finally, to reduce computational load, this model stratified the population into four age groups, with the 5-64-year-old age group comprising the majority of the population. Since the model did not include restricted carriage transmission from adults to children, this age stratification may result in an under-estimation of disease among all age groups. Finally, future work should consider estimating the public health effects associated with other higher valency pediatric vaccines, including a 20-valent PCV (PCV20).

## CONCLUSIONS

Switching from PCV13 to PCV15 for routine pediatric vaccination via the 1+1 dosing schedule in the UK would not only further reduce IPD in the pediatric population but would also lead to population-level reductions in IPD due to indirect protection. Although this model predicted short-term increases in VT-IPD associated with potential reductions in VE against disease in a reduced dosing schedule, the effects were not persistent into the future, and these transient increases were smaller in magnitude with the implementation of PCV15 than continued use of PCV13. In countries with lower pediatric VCRs, nascent pneumococcal vaccination programs, or uncontrolled VT-IPD, a reduced dosing schedule may lead to more pronounced and persistent increases in VT-IPD.

## Data Availability

All relevant data are within the manuscript and its Supporting Information files. This is a modeling study and, therefore, no primary data was collected in this study. All inputs were from published literature and included only anonymized data.

## ACKNOWLEDGEMENTS

We wish to thank UKHSA for providing seroprevalence data, and we wish to thank Kevin Bakker and Kenneth Klinker for their efforts in the development of the UK model adaptation. We also wish to thank Robert Nachbar for his useful comments and suggestions during model development and adaptation to the UK, and Colleen Burgess for her assistance in the preparation of this manuscript.

## Disclosures

RJO, NB, JW, TMM, and OS are employees of Merck Sharp & Dohme LLC, a subsidiary of Merck & Co., Inc., Rahway, NJ, USA. IRM and DN are employees of MSD (UK) Ltd., London, United Kingdom. GM is an employee of Wolfram Research Inc. under contract to Merck Sharp & Dohme LLC, a subsidiary of Merck & Co., Inc., Rahway, NJ, USA. JCL is an employee of Merck Canada, Inc., Kirkland, QC, Canada. RJO, NB, JW, IRM, DN, TMM, JCL, and OS may hold stock or stock options in Merck & Co., Inc., Rahway, NJ, USA.

## Funding

This study was funded by Merck Sharp & Dohme LLC, a subsidiary of Merck & Co., Inc., Rahway, NJ, USA. The funder provided support in the form of salaries or consulting fees for authors.

## SUPPLEMENTARY FIGURES / TABLES

**Figure S1.**
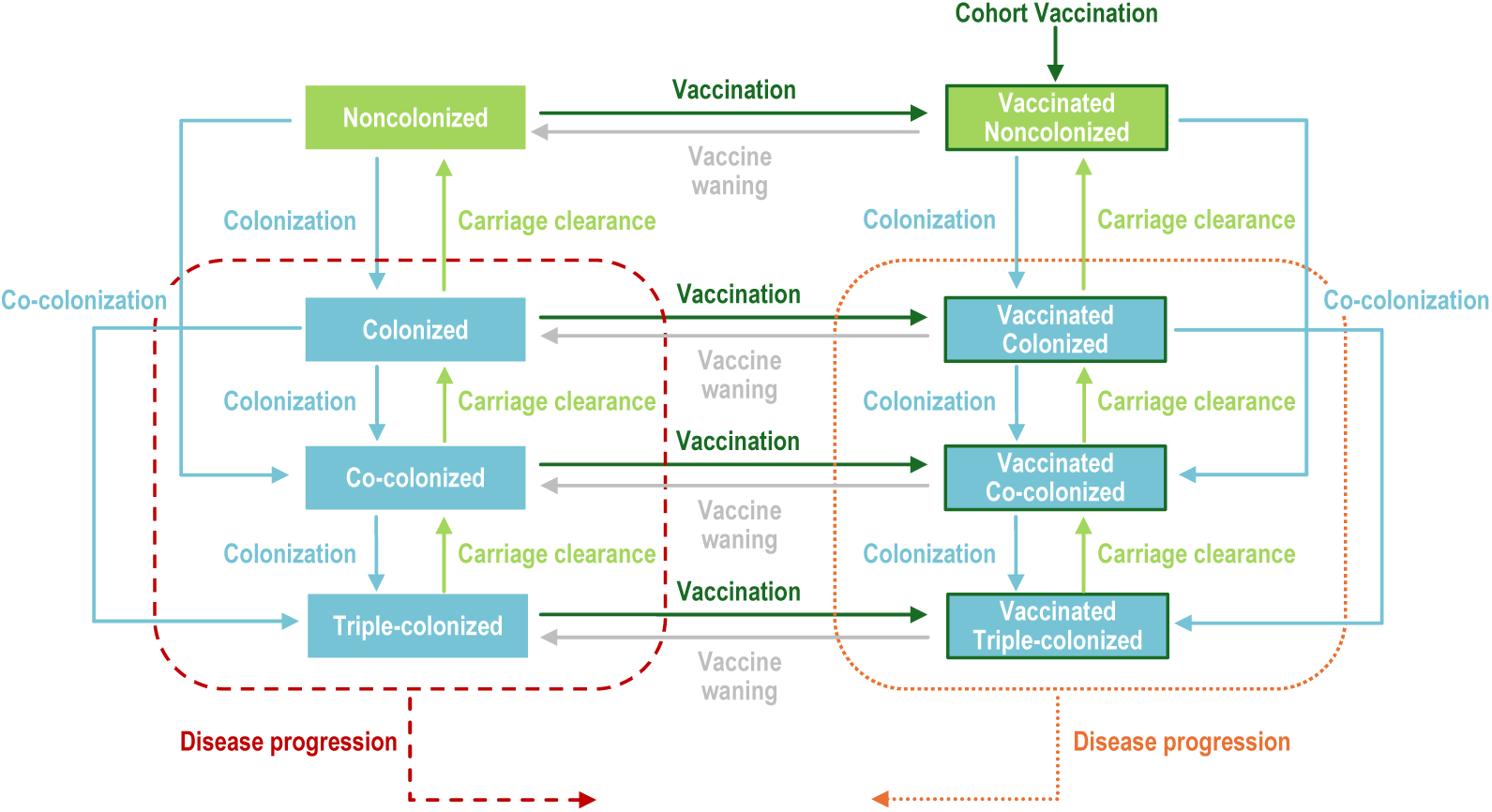
Model diagram. Upon carriage acquisition, individuals moved to either the colonized, co-colonized, or triple-colonized classes, depending on whether one or two serotypes were acquired (denoted in blue). Some portion of individuals from these colonized classes would develop a pneumococcal disease (denoted in red and orange). Recovery then moved individuals back to the non-colonized class (denoted in light green), sequentially clearing one serotype at a time. Vaccination (dark green border) reduced the risk of carriage acquisition and disease development for vaccine serotypes.

**Figure S2.**
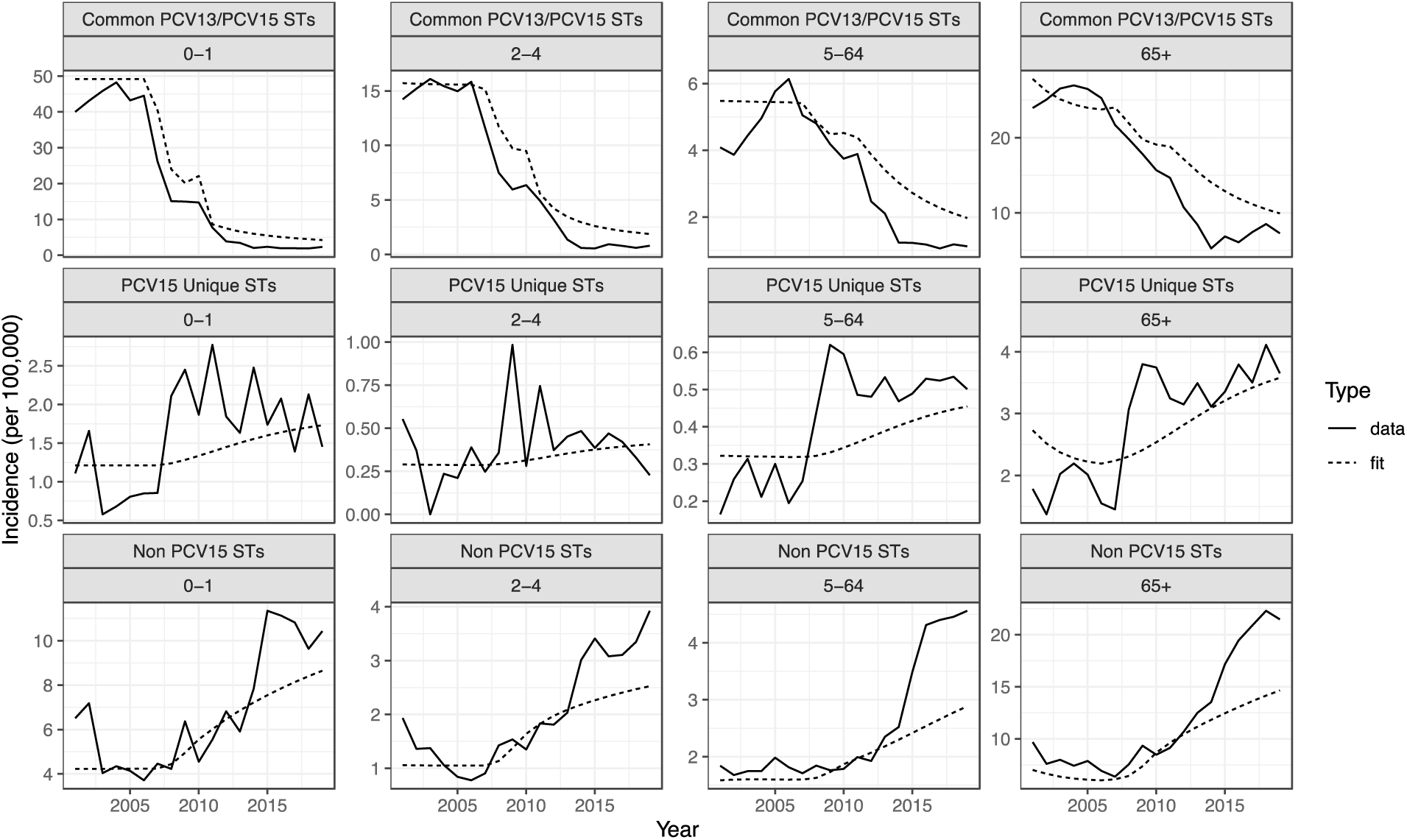
Model calibration to historical observed IPD incidence per 100,000 population by ST grouping and age group. Comparison of data (solid line) to model fit (dotted line) during the calibration period faceted by age group and vaccination grouping.

**Figure S3.**
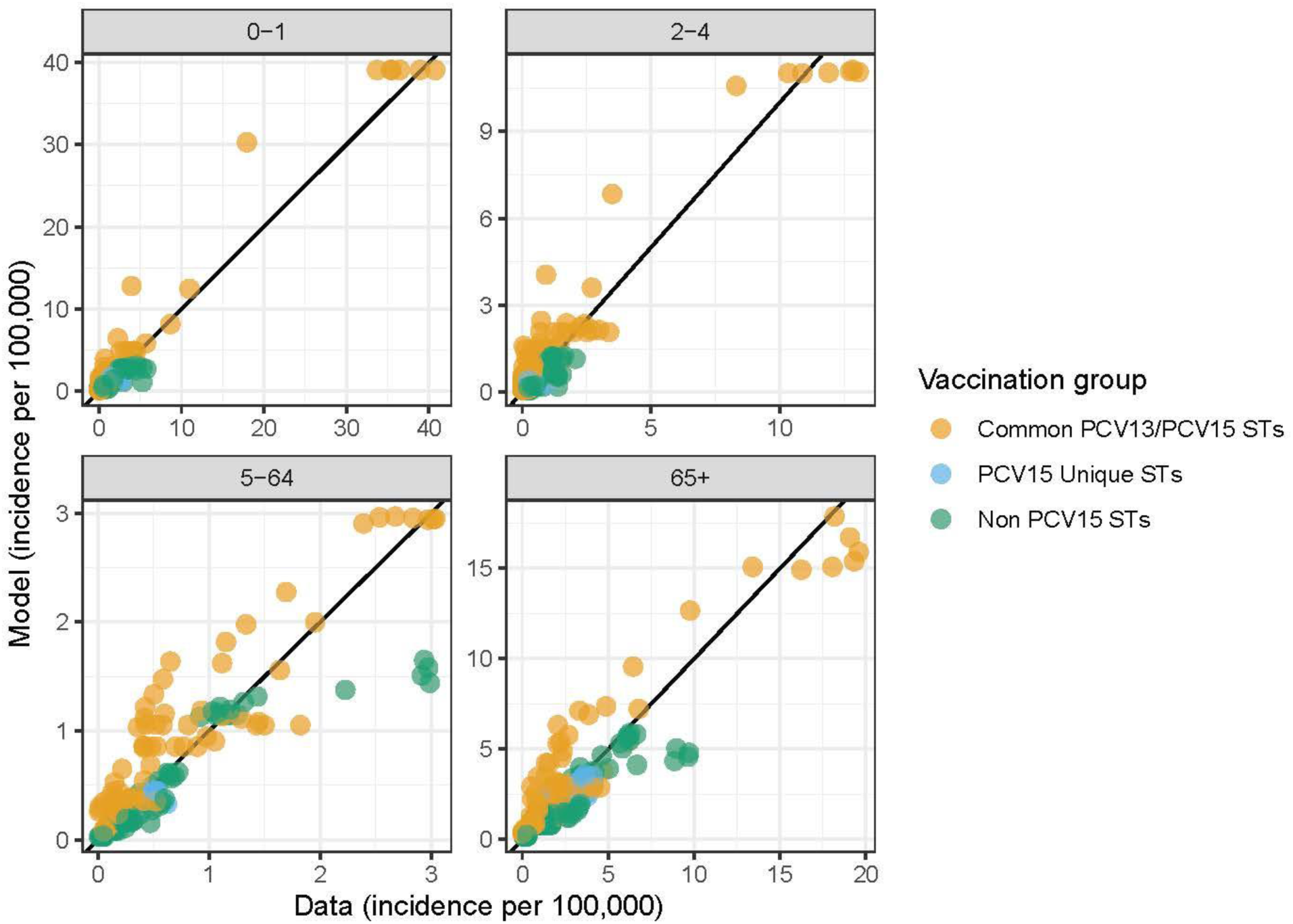
Model validation of calibrated model comparing data to model fit during the calibration period, by age group. Points denote annual and STC-specific incidence and are colored by vaccination grouping. Black line indicates a 1:1 line. Root mean squared error (RMSE) values were 1.47, 0.597, 0.312, and 1.29 for the <2-, 2-4-, 5-64-, and ≥65-year-old age groups. Across all age groups, the RMSE was 1.03.

---

**Table S1.** Vaccine efficacy against disease for the <2-year-old age group ^22^ for a 2+1 schedule.

| Serotype class (STs) | VE against disease (PCV7) | VE against disease (PCV13) | VE against disease (PCV15) |
| --- | --- | --- | --- |
| 1 (4, 6B, 9V, 14, 18C, 19F, 23F) | 0.96 | 0.96 | 0.96 |
| 2 (1, 5) | 0 | 0.84 | 0.84 |
| 3 (3) | 0 | 0.41 | 0.41 |
| 4 (7F, 19A) | 0 | 0.93 | 0.93 |
| 5 (6A, 6C) | 0 | 0.96 | 0.96 |
| 6 (22F, 33F) | 0 | 0 | 0.94 |
| 7 (9N, 17F, 20) | 0 | 0 | 0 |
| 8 (8, 10A, 11A, 12F) | 0 | 0 | 0 |
| 9 (15B) | 0 | 0 | 0 |
| 10 (15A, 15C, 16F, 23A, 23B, 24F, 31, 35B) | 0 | 0 | 0 |
| 11 (NVTs) | 0 | 0 | 0 |

VE = vaccine efficacy; NVTs = non-vaccine types; STs = serotypes

**Table S2.** Vaccine efficacy against disease for the 2-4-year-old and 5-64-year-old risk group and ≥65-year-old population for PPSV23. ^23^ Note that given the lack of real-world evidence for vaccine effectiveness against disease for novel vaccine serotypes (STCs 6-10), we assumed equivalency with previous VTs in adult PCVs.

| Serotype class (STs) | VE against disease (PPSV23) |
| --- | --- |
| 1 (4, 6B, 9V, 14, 18C, 19F, 23F) | 0.45 |
| 2 (1, 5) | 0.23 |
| 3 (3) | 0.22 |
| 4 (7F, 19A) | 0.59 |
| 5 (6A, 6C) | 0 |
| 6 (22F, 33F) | 0.54 |
| 7 (9N, 17F, 20) | 0.65 |
| 8 (8, 10A, 11A, 12F) | 0.57 |
| 9 (15B) | 0.53 |
| 10 (15A, 15C, 16F, 23A, 23B, 24F, 31, 35B) | 0 |
| 11 (NVTs) | 0 |

VE = vaccine efficacy; NVTs = non-vaccine types

**Table S3.**
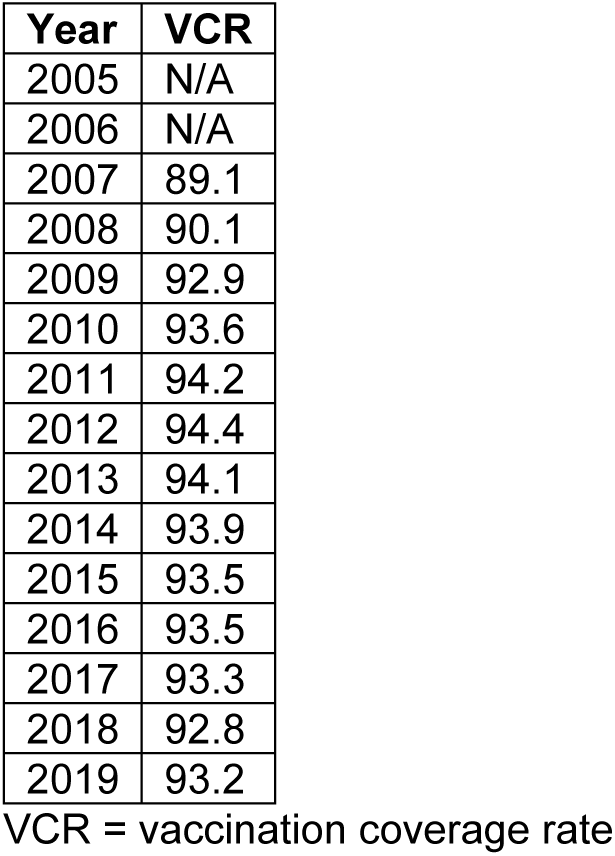
PCV Coverage for the <2-year-old age group. ^39^

**Table S4.**
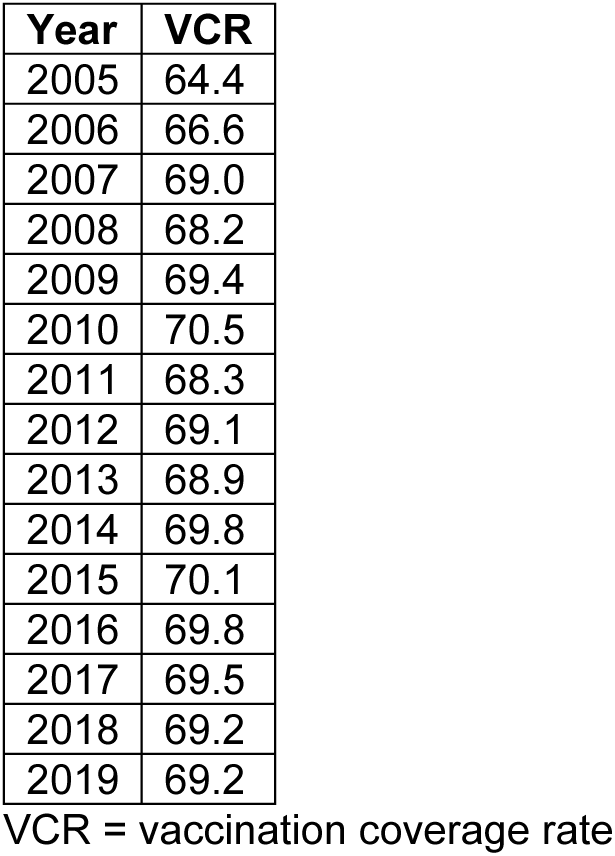
PPSV23 Coverage for the ≥65-year-old age group. . Data were from 2005-2018. For 2019, we assumed the same coverage as 2018.^40^

| Year | VCR |
| --- | --- |
| 2005 | 64.4 |
| 2006 | 66.6 |
| 2007 | 69.0 |
| 2008 | 68.2 |
| 2009 | 69.4 |
| 2010 | 70.5 |
| 2011 | 68.3 |
| 2012 | 69.1 |
| 2013 | 68.9 |
| 2014 | 69.8 |
| 2015 | 70.1 |
| 2016 | 69.8 |
| 2017 | 69.5 |
| 2018 | 69.2 |
| 2019 | 69.2 |

VCR = vaccination coverage rate

**Table S5.**
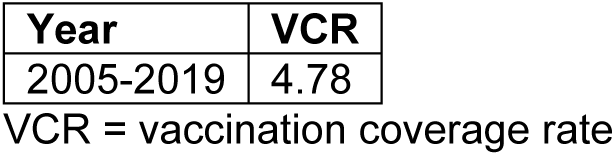
PPSV23 Coverage for the at-risk 2-4- and 5-64-year-old age group. These calculations assume that the VCR of clinical risk groups is 49.0% and that the proportion of the 2-64-year-old population that falls within a clinical risk group is 9.7%.^41^

